# Neonatal apnea and hypopnea prediction in infants with Robin sequence with neural additive models for time series

**DOI:** 10.1101/2023.03.14.23287021

**Authors:** Julius Vetter, Kathleen Lim, Tjeerd M. H. Dijkstra, Peter A. Dargaville, Oliver Kohlbacher, Jakob H. Macke, Christian F. Poets

**Affiliations:** Machine Learning in Science, Tübingen University and Tübingen AI Center, Tübingen, Germany; Department of Computer Science, University of Tübingen, Germany; Menzies Institute for Medical Research, College of Health and Medicine, University of Tasmania, Hobart, Tasmania, Australia; Institute for Translational Bioinformatics, University Hospital Tübingen, Germany; Department of Women’s Health, University Hospital Tübingen, Germany; Neonatal and Pediatric Intensive Care Unit, Department of Pediatrics, Royal Hobart Hospital, Hobart, Tasmania, Australia; Institute for Bioinformatics and Medical Informatics, University of Tübingen, Germany; Max Planck Institute for Intelligent Systems, Tübingen, Germany; Department of Neonatology, University Hospital Tübingen, Germany

## Abstract

Neonatal apneas and hypopneas present a serious risk for healthy infant development. Treating these adverse events requires frequent manual stimulation by skilled personnel, which can lead to alert fatigue. Automatically predicting these adverse events before they occur would enable the use of methods for automatic intervention. In this work, we propose a neural additive model to predict individual events of neonatal apnea and hypopnea and apply it to a physiological dataset from infants with Robin sequence at risk of upper airway obstruction. The dataset will be made publicly available together with this study. Our model achieved an average area under the receiver operating characteristic curve of 0.80 by additively combining information from different modalities of the respiratory polygraphy recording. This permits the prediction of individual apneas and hypopneas up to 15 seconds before they occur. Its additive nature makes the model inherently interpretable, which allowed insights into how important a given signal modality is for prediction and which patterns in the signal are discriminative. For our problem of predicting apneas and hyponeas in infants with Robin sequence, prior irregularities in breathing-related modalities as well as decreases in SpO_2_ levels were especially discriminative.

## Introduction

It would be of high clinical significance to predict apnea and hypopnea events in newborn infants before they occur, in order to perform preventative automated intervention. Automatic intervention could, for example, be realised by an inflatable mattress through which stochastic vibrotactile stimulation can be applied [1].

Compared to the amount of work in automatic apnea *detection* [2, 3, 4, 5], there has been considerably less work in automatic apnea and hypopnea *prediction* [6]. Prior work on prediction has mainly focused on associated events of bradycardia and used cardio-respiratory features together with different classifiers like hierarchical classification methods [7] and random forests [8]. Recent studies also used infant movement as an additional feature [6]. Another study more generally used deep neural networks to identify infants more susceptible to apnea and hypopnea [9].

However, previous work did not address predicting individual events of apnea or hypopnea, but rather tried to predict whole episodes with repeated apneas and hypopneas [6]. Consequently, the prediction horizon for the cited approaches was on the order of several minutes [10].

In this study, we were instead interested in predicting individual events of apnea and hypopnea and thus worked on a time scale of seconds. Because of this new time scale, there were no easily available or traditionally used features to extract from the recorded signals. In recent years, deep neural networks have had considerable success in automating the process of feature extraction. Many fields of science, including the prediction of adverse events in medical time series, have profited from this progress [11, 12, 13]. However, a major drawback of classical deep neural network architectures is that they are a black box: Because of their complexity, they make it difficult to understand why a certain prediction was made and which features of the signals were contributing. Especially in the medical domain, where mistakes are costly, opening this black box and making its decisions more interpretable is crucial for applications. There exist several methods to generate post-hoc explanations of black-box models, but their use was recently discouraged in high-risk applications due to unreliable or misleading explanations [14]. An alternative to post-hoc explanations are models that are interpretable by construction. Creating such models is often possible without a drop in model performance [15].

### Our Contribution

We built on recent work in neural additive models [16], which form their prediction by summing over the output of several neural networks. The additive nature allows users to inspect the magnitude of the additive contributions for each prediction to gauge the importance of the underlying feature or signal modality. This inherent interpretability makes neural additive models a good choice for clinical prediction tasks like individual apnea or hypopnea prediction. Since our data came in the form of time series, we replaced classically-used neural network architectures by architectures which are tailored to time series. We show that our model achieved a high level of prediction, with an average area under curve of 0.80 for the task of apnea and hypopnea prediction for infants with Robin sequence. Furthermore, our model allowed us to gauge the importance of different signal modalities for a performed prediction as well as to localise discriminative features within the signals.

## Results

### Neonatal dataset and problem setup

We performed our analyses on a dataset of *n* = 19 infants with Robin sequence who had been admitted to the Tübingen neonatology department between May 2020 and April 2021. Gestational age at birth was 39 (32–41) [median (range)] weeks and birth weight 3,390 (1,320–4,380) g. The recording methods are described in detail in [17]. At the time of conducting the respiratory polygraphy, infants were 17 (1–73) days old and weighed 3,392 (2,642–4,380) g. In total, 185 hours of respiratory polygraphy data were recorded, containing 122 hours of total sleep time. Adverse events, namely obstructive, mixed and central apneas, and hypopneas, as well as infant movement were annotated via visual inspection by a domain expert after the recording. Infants experienced 27 (3–112) obstructive, mixed and central apnea events per hour, and 28 (1–58) obstructive and mixed hypopnea events per hour, with events lasting for 3.8 (3.9–5.7) and 4.9 (2.8–7.3) seconds per event, respectively. After removing parts of the signals that were annotated as either adverse events or movements, signals were divided into 30 second time windows. The target time windows were taken with an offset of 15 seconds, whereas the control time windows were required to be at least three minutes away from both the start and end of any adverse event. This procedure resulted in 214 (75-606) time windows per infant to be used for classification. For most patients there were more control than target time windows. The imbalance (percentage of target time windows) was 36% (4%-66%). The overall setup is depicted in Fig. 1 and is described in more detail in the Method Section.

**Figure 1:**
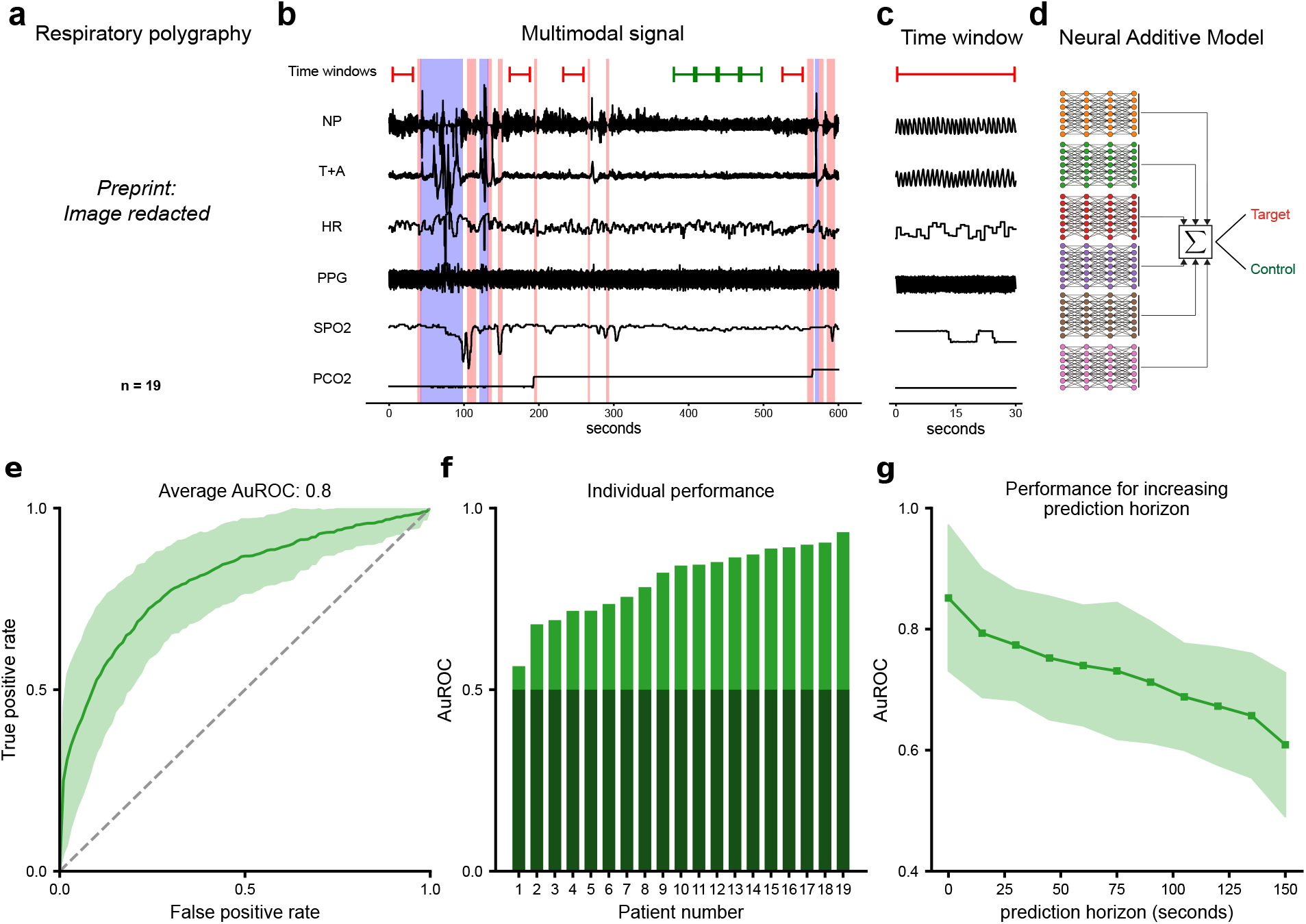
Overview of the setup and core results. a) 19 overnight respiratory polygraph recordings of infants with Robin sequence were collected. Six signals were used for the analysis: Nasal pressure (NP), the sum of thoracic and abdominal respiratory effort (T+A), photoplethysmogram (PPG), heart rate (HR), SpO_2_, and PCO_2_ levels. b) Adverse events (apneas and hypopneas, indicated as pink vertical boxes) as well as infant movement (blue boxes) were annotated manually. From these annotations, 30 second target time windows shortly preceding the adverse events (red), as well as control time windows (green) were extracted. c) Example of an extracted target time window. d) To discriminate between target and control time windows, a Neural Additive Model (NAM) [16] was trained and tested with patient-based leave-one-out cross validation. e) Classification performance of the NAM: Pointwise average over the ROC curves of individual patients. Uncertainty corresponds to the pointwise standard deviation. f) Area under ROC values for the individual patient-based test sets sorted in increasing order. Random performance is indicated in dark green. g) The average test performance as a function of an increasing prediction horizon. Uncertainty corresponds to the standard deviation over the performance of different patients.

### Performance of neonatal adverse event prediction

We constructed a time-series based Neural Additive Model (NAM) [16] to perform the classification into target (pre-adverse event) and control time windows based on seven signal modalities: Nasal pressure, thoracic and abdominal respiratory effort, heart rate, SpO_2_, and PCO_2_ levels. Because of their strong correlation, thoracic and abdominal respiratory effort were summed to obtain a single signal, resulting in six modalities. We measured the performance of our classifier by performing patient-based leave-one-out cross validation. The average area under the receiver operating characteristic curve (AuROC) over all leave-one-out test sets was 0.80 with a standard deviation of 0.093 (Fig. 1). For some patients, we achieved a classification performance of over 0.9 AuROC. We conducted permutation tests [18] to assess that our obtained test performances are significant. For all but one patient (*p*_*per*_ = 0.121), the performance was significantly better than random (*p*_*per*_ *<* 0.001).

We also investigated how much performance drops as the prediction horizon between target time window and adverse event is increased or decreased. Fig. 1f shows the average AuROC for completely retrained classifiers as a function of the prediction horizon. As expected, for an increasing prediction horizon, the classification performance decreased and reached near chance level after about 150 seconds. On the other hand, for no offset between annotated events and target time windows, the performance further increased.

### Neural Additive Model for signal modality importance

Our neural additive model allowed us to investigate how predictions are formed by inspecting the corresponding additive contributions. A key question was to understand which of the six physiological signal modalities carry predictive information. Identifying which signals carry the most predictive information could enable both neonatologists and possible future automated prediction systems to focus on fewer signal modalities, thus reducing strain on the infants. The NAM classifier allowed us to investigate the importance of different signal modalities by analysing the additive contribution of each signal modality to the overall classification score. This analysis was possible because, by construction, the overall classification score is a sum of the individual additive contributions from each signal modality. The higher an individual additive contribution is compared to the other contributions, the more influence it has on the overall classification. To gauge overall importance of the six signal modalities used for classification, we analysed the additive contributions pooled over all patient test sets (Fig. 2a). To analyse differences between patients, we also computed the standard deviations over the additive contributions of individual patients (Fig. 2b). Furthermore, we performed a single modality analysis, where the subnetwork of each signal modality is trained independently. To analyse the significance of potential differences between modalities, we repeated our training procedure five times and computed Wilcoxon signed-rank (*p*_*wil*_) tests over the *n* = 19 patients. See the Appendix for more details about the statistical tests.

**Figure 2:**
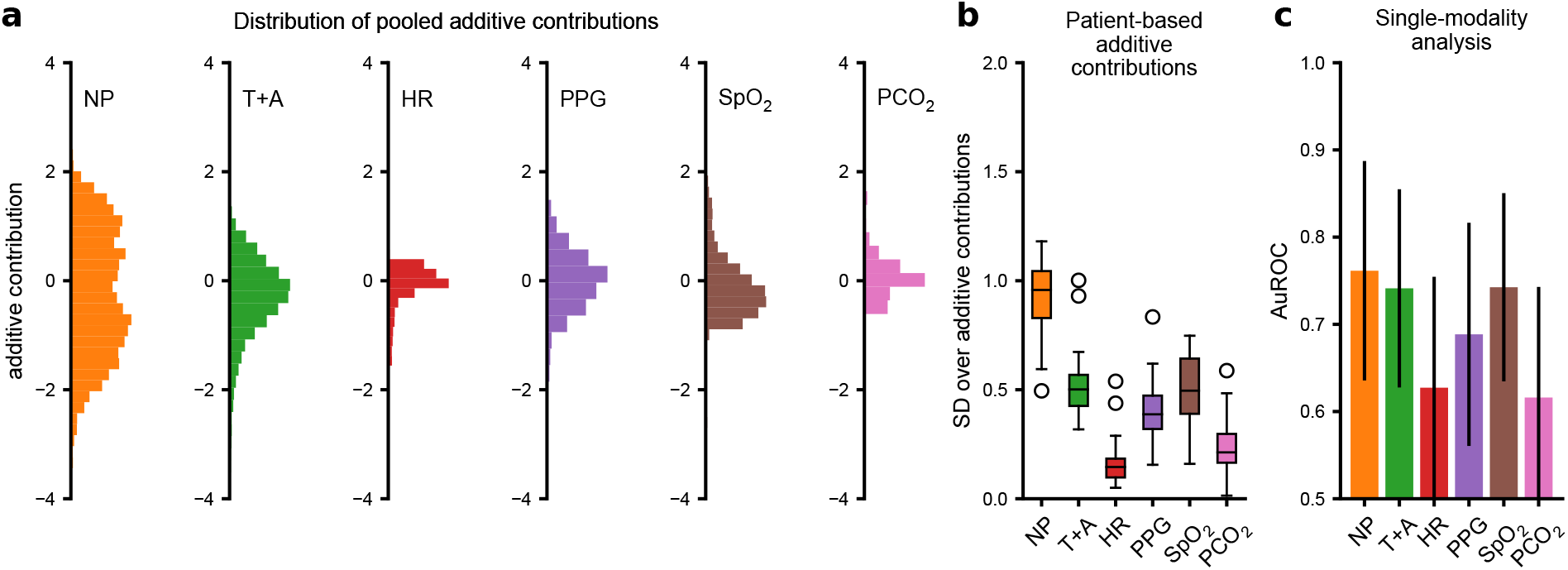
Importance analysis of the signal modalities. For the NAM, the importance of a signal modality can be measured by analysing its associated additive contributions. a) Histograms of additive contributions pooled over all patient-based test sets. b) Box plots of the distribution over the standard deviation of additive contributions of all individual patients. c) Average AuROC scores of the individually trained single modality networks together with the standard deviation. All analyses show that the nasal pressure signal contains the most predictive information on average.

**Figure 3:**
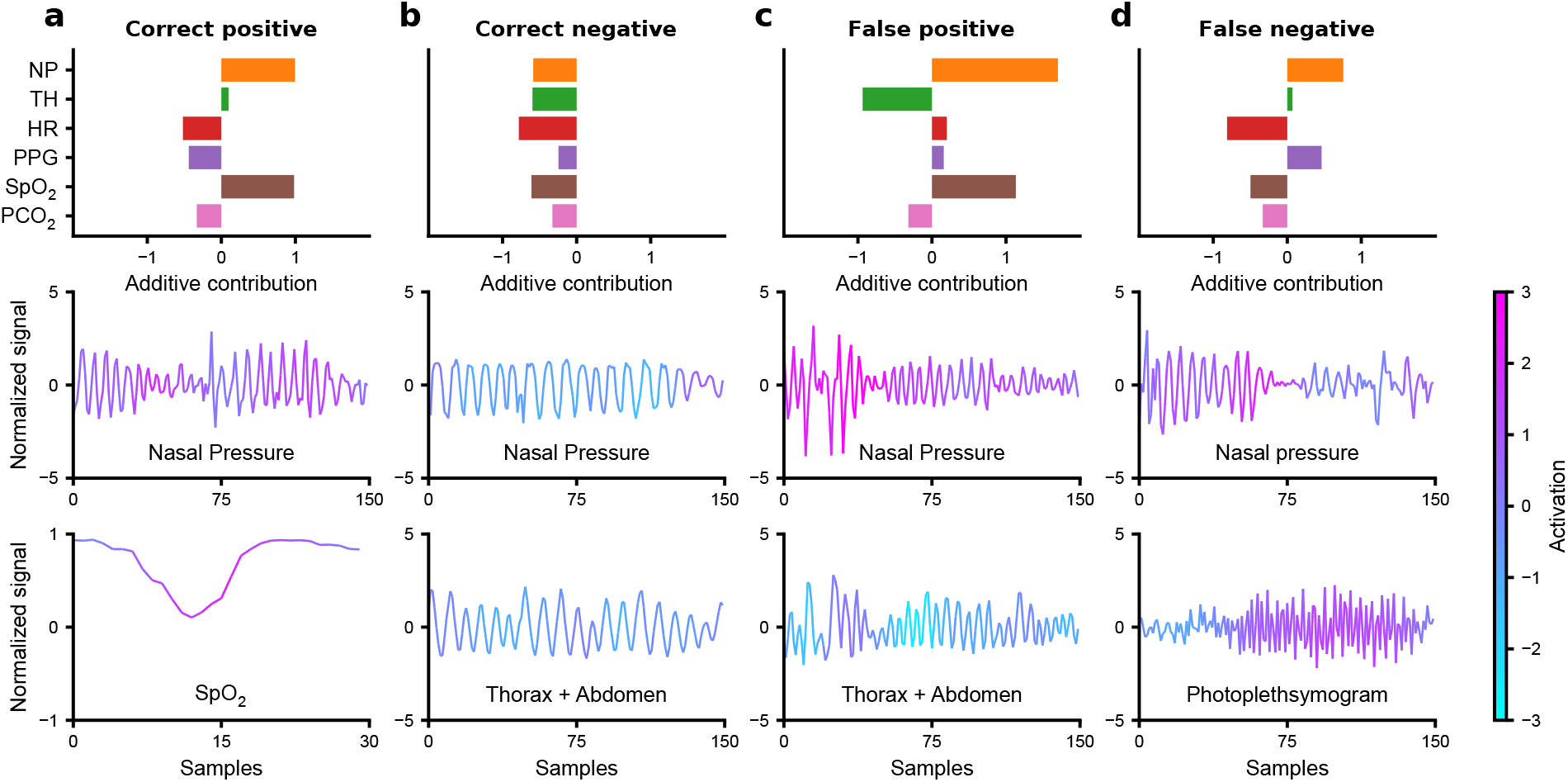
Exemplary activation maps together with the associated additive contributions. a) Correctly classified target (pre-adverse-event) time window. Light blue indicates negative activation and a classification towards no adverse event. Pink indicates positive activation and a classification towards an upcoming adverse event. The classifier detected different types of irregularities in the breathing signal as well as variations in heart rate and SpO_2_ levels. b) Correctly classified control time window c) Incorrectly classified control time window (false positive). d) Incorrectly classified target time window (false negative).

The large standard deviations over both the pooled and patient based additive contributions indicated that the nasal pressure signal contained the most predictive information (Fig. 2a/b). The other breathing-related signal, the sum of thoracic and abdominal respiratory effort, was comparatively less informative (*p*_*wil*_ *<* 0.001). This result was confirmed by the performances of the single modality networks: The network for nasal pressure achieved the highest average AuROC, which was significantly better than the performance achieved by the network for thoracic and abdominal respiratory effort (*p*_*wil*_ = 0.032) (Fig. 2c). The importance of both breathing-related signals was in accordance with the clinical perspective as infants with Robin sequence mainly suffer from obstructive apnea and hypopnea. Furthermore, both the NAM and single modality analysis showed that, apart from the breathing-related signals, the other modalities PPG, heart rate, PCO_2_ and especially SpO_2_ also carried predictive information. The SpO_2_ single modality performance was on par with the nasal pressure (no significant difference: *p*_*wil*_ = 0.241). The informativeness of SpO_2_ was expected clinically: apneas and hypopneas often appear in clusters and hence decreased SpO_2_ levels may indicate immediately preceding adverse events which increase the probability of another upcoming adverse event. A similar effect could be observed for the heart rate as apneas and hypopneas are often followed by bradycardia. Furthermore, the PPG signal is very sensitive to small movements, and thus captured pre-apnea or pre-hypopnea arousal.

Although some of the signal modalities contained less information than others, additively combining their information increased overall performance: The NAM achieved significantly better average AuROC than each of the trained single modality networks (*p*_*wil*_ = 0.009).

### Activation maps for local interpretability

While analysing the additive contributions of the NAM revealed which of the signal modalities contained the most predictive information, it could not answer what parts or features of the respective signal modalities were discriminative for an upcoming event. In other words, we wanted to answer the question: What does the classifier “look at” within the signal modalities? As we input whole time series into our classifier, we lost global interpretability, that is, the ability to visualise the full regression function, which is usually provided in generalised additive models. We could, however, achieve local interpretability. Local interpretability refers to the the explanation for the prediction made with respect to an individual data point. To this end, the fully convolutional subnetworks of the NAM allowed us to use convolutional activation maps to investigate which part of the time window activates the convolutional filters.

In our case, each time window classified by the NAM resulted in a total of six activation maps and additive contributions, one for each signal modality. Additive contributions were simply computed as the average activation across time. Positive activations indicated that the corresponding segment of the signal was discriminative towards a target time window. In contrast, negative activations indicated that a segment of the signal was discriminative towards a control time window. Based on a visual analysis, the classifier focused on irregularities in the breathing signal both for the nasal pressure and the sum of the thoracic and abdominal respiratory efforts. The detected irregularities in the nasal pressure could include general change in respiratory rate and amplitude or a “mini-apnea” (flat nasal pressure signal not meeting annotation criteria). On top of these features, the classifier used arousal in the plethysmograph, reduced SpO_2_ levels or SpO_2_ desaturations as well as variations in the heart rate to make a classification towards a pre-adverse-event time window.

## Discussion

We developed an interpretable time-series classifier and applied it to the problem of neonatal apnea and hypopnea prediction in infants with upper airway obstruction. Unlike previous work, we focused on predicting individual events of apnea and hypopnea. Our neural additive model (NAM) was able to automatically extract relevant features from the multimodal polygraphy signal and achieved good performance in classifying individual time windows. Importantly, because of the inherent interpretability of the NAM, we were able to perform several downstream analyses to provide insights into the features relevant for individual apnea and hypopnea prediction.

First, the NAM allowed us to investigate the importance of each signal modality recorded with respiratory polygraphy. Instead of the classically-used thoracic (and abdominal) respiratory effort [6], the nasal pressure turned out to be the more informative breathing-related modality in predicting individual events. Apart from breathing-related modalities, other modalities were also informative, although to a lesser degree. Nevertheless, their inclusion into the NAM resulted in a superior performance compared to the individual single-modality networks. From a practical point of view, the predictive importance of nasal pressure potentially allows practitioners to reduce the burden of respiratory polygraphy. If full respiratory polygraphy is not required for scientific or clinical reasons, overnight monitoring might be possible with only the nasal pressure.

Second, with the help of activation maps, we were able to visualise the learned features. These visualisations allowed us to confirm their clinical significance. The activation maps could also play an important role in future clinical applications, where nurses and neonatal intensive care unit (NICU) staff work together with automated systems for apnea and hypopnea prediction. Based on the local importance indications provided by the activation maps, nurses and NICU staff could decide whether to trust or distrust the prediction made by the model. In case of systematic false alarms, NICU staff could then intervene by recalibrating the model or, in extreme cases, turning it off altogether. Allowing this type of distrust in a model is crucial for application in real-world clinical settings [14].

### Limitations

While this study presents a first step in the direction of individual neonatal apnea and hypopnea prediction, there are several obstacles that hinder the application of the presented model in a real-world clinical setting. The ultimate goal would be to create a system that monitors an infant, predicts adverse events, and automatically triggers an intervention (that is, a closed loop apnea management system).

The presented model is not yet well suited for this task. For some patients, the prediction performance was not sufficient to be of practical use. Especially for systems that perform automated interventions, high prediction accuracy will be necessary as mistakes in the clinical setting are costly. Note that we evaluated performance by training and validating on 18 of the 19 infants and testing on the one left out. Following a domain adaptation approach, where the classifier adapts to the current infant in an online manner, could potentially mitigate the low performance for some of the neonates. Furthermore, additional measures would need to be taken to ensure the robustness of the model to unseen data, outliers and malfunctioning recording equipment to guarantee safe deployment.

There is also room for improvement regarding the interpretability of the classifier. As presented, the classifier did not discriminate between different predictive features within the time series that occur before an adverse event. A model that can provide more context about these predictive features will further increase the trust in its ability to provide correct predictions.

More generally, the current NAM classifier ignores possible interactions between time series. This shortcoming, common to all generalized additive models (GAMs), can lower the model expressiveness and cause concurvity issues that negatively affect the interpretability and stability of the model. There is work on including two-feature interaction in tabular GAMs [19], but it is currently unclear how to extend this to the time-series based NAM framework without losing some of the inherent interpretability.

### Conclusion

Despite its limitations, our neural additive model presents a first step towards automated neonatal apnea and hypopnea prediction. When performing reliably, such a model would have the potential to optimise care for vulnerable neonates with upper airway obstruction.

## Methods

### Data collection

We based our study on a physiological dataset from infants with Robin sequence. This group is well suited for automated prediction because of their homogeneous pathophysiology, that is, apneas and hypopneas are mainly caused by a narrow upper airway. Between May 2020 and April 2021, 19 infants with Robin sequence underwent whole night recordings in the Department of Neonatology at Tübingen University Hospital, using standard digital cardiorespiratory polysomnography (Remlogic, Natus Medical Incorporated, California, USA). Modalities recorded included nasal airflow obtained via a nasal pressure transducer and recorded at 200Hz, as well as thoracic and abdominal respiratory efforts via respiratory inductance plethysmography recorded at 50Hz. Furthermore, the heart was monitored via an EKG and PPG recorded at 200Hz and 100Hz respectively. The beat-to-beat heart rate was automatically derived from the EKG signal based on the RR-intervals. Finally, the blood SpO_2_ obtained from pulse oximetry and transcutaneous PCO_2_ readings were recorded. Manual annotations of adverse events were derived from the recordings [17]. Five different types of adverse events were annotated by a domain expert according to the criteria of the 2020 American Academy of Sleep Medicine (AASM) guidelines. These were central apnea, obstructive apnea, and mixed apnea as well as central and obstructive hypopnea. Additionally, intermittent hypoxia events, defined as a fall in SpO_2_ levels by more than three percentage points within a duration of five seconds (desaturations) and body movements were annotated. The movement annotation was based on a low-resolution infrared video frame. Details about the annotation criteria can be found in the Appendix.

### General Setup

We framed the problem of apnea and hypopnea prediction as a binary time-series classification problem. All types of apnea and hypopnea were combined into one single type of adverse event. We made no attempt to differentiate between different types of apneas and hypopneas. Intermittent hypoxia events were not considered as adverse events. Parts of the signal that were annotated as either an adverse event or movement were removed from the signal. The remaining signal was then divided into 30 second non-overlapping time windows. If a time window preceded an adverse event with an offset of 15 seconds, it was labeled as a target time window. This offset was well above the maximal duration of the annotated apneas and hypopneas, and thus ensured that our prediction was not based on imprecise annotation time stamps. If a time window was at least three minutes away from both the start and end of an adverse event, it was labeled as a control time window (Fig. 1).

As mentioned in the Results Section, the number of extracted time windows ranged between 75 and 606 time windows with a median number of 214. For all but one patient, the time window generation resulted in more control time windows than pre-adverse-event time windows. The imbalance ranged between 4% and 66% with a median imbalance of 36%. To avoid training issues due to class imbalance, we undersampled the control time windows on a patient basis during training to balance control and target time windows. The classifier performance was evaluated on the full (imbalanced) test sets. To assess the average performance of our classifier, we performed patient-based leave-one-out cross validation. That is, we used all generated time windows from a single patient as a test set and trained on the union of all time windows of the remaining patients. This procedure was repeated for every patient and average performance scores and standard deviations are reported. For every repetition, we selected the hyperparameters with nested cross validation [20]: All time windows from every patient apart from the test patient time windows were used as a validation set. After exploring different hyperparameter configurations, we picked the hyperparameters that resulted in the highest average validation AuROC for training. More details are given in the Section “Architecture and training details”.

We focused on the importance of six signal modalities: Nasal pressure, the sum of thoracic and abdominal respiratory effort, heart rate (which is derived from the ECG), PPG, SpO_2_, and PCO_2_ signals. Inter-breath intervals based on thoracic respiratory effort and heart rate have “classically” been used to predict apnea and hypopnea [6]. Because thoracic and abdominal respiratory effort are highly correlated signals, we treated them as a single signal by summing both signals traces pointwise. Creating the sum of these two modalities is a well-established practice in the analysis of apnea and hypopnea episodes [21, 22]. Additionally, nasal pressure presented another interesting breathing-related signal that potentially carries different information than the “classically” used thoracic and abdominal respiratory effort. Moreover, we included the PPG signal and the two blood related parameters SpO_2_ and PCO_2_.

### Preprocessing

The three breathing-related signals nasal pressure, thoracic and abdominal respiratory effort, sampled at 200Hz or 50 Hz respectively, were downsampled to 5 Hz after applying an anti-aliasing 8th order Chebyshev filter. The same preprocessing was applied to the PPG signal. Both the SpO_2_ and PCO_2_ signals were downsampled from 2Hz to 1Hz. The derived heart rate was left unchanged at 1Hz. We then standardised all breathing-related signals and the PPG signal on a time window basis. The summed signal of the thoracic and abdominal respiratory effort was obtained by summing both standardised signals pointwise and standardising the result again. The heart rate as well as SpO_2_ and PCO_2_ were range-normalised from 50 to 240bpm, 60-100%, and 30-70 mmHg to a range of -1 to 1. Our rationale for standardising the breathing-related and PPG modalities but range-normalising the heart rate, SpO_2_ and PCO_2_ signals, was that signal values from first group of modalities can vary strongly between patients. This is not true for the latter group of modalities. Apart from downsampling and standardising or normalising, we performed no additional preprocessing or filtering, allowing the capture of systematic noise in the signals, which might be predictive for an upcoming adverse event.

### Periodic breathing

The choice of 30 second time windows was not arbitrary. Since we did not perform padding for shorter time windows, the fixed length ensured a minimal distance between annotated adverse events. This fixed length was necessary to avoid the very short inter-apnea episodes that occur during periodic breathing and that are easily distinguishable from normal breathing [23, 6]. With the choice of 30 seconds, we were above the definition of Kelly and Shannon [24], who define periodic breathing as “three or more episodes of central apnea lasting at least 4 seconds, each separated by no more than 20 seconds of normal breathing”.

### Interpretability

In medical adverse event prediction, there are usually several signal modalities available that might be used to predict adverse events. Furthermore, data is often noisy and heterogeneous. It is therefore important to have some level of interpretability to avoid unforeseen shortcuts and failure modes in the prediction model. Our case of neonatal apnea and hypopnea prediction was no different in this regard. Since there was little prior work on possible features for individual apnea and hypopnea prediction [10], we performed automatic feature extraction with deep neural networks. Despite using deep neural networks, we required our classifier to be inherently interpretable. There were two questions we wanted our classifier to be able to answer:

- Question 1: Which physiological signal modalities of the multivariate polygraphy signal carry the strongest information to make a prediction?
- Question 2: What features within the signal modality traces themselves are discriminative for the prediction?

The first question could be answered by inspecting the additive contributions of a prediction made by a neural additive model and the second by generating activation maps. Both approaches were easily unified into a single architecture, as we describe in the following.

### Neural Additive Models (Question 1)

Generalised Additive Models (GAMs) have historically been used to analyse tabular data especially in the medical domain. They combine expressiveness with built in interpretability [25, 19, 26]. For tabular data, GAMs are a linear combination of non-linear functions *f*_*i*_, which each take one feature of the data as input. Linear and logistic regression are two special cases of GAMs where the *f*_*i*_ are constrained to be linear functions. The basic GAM for binary classification is given by

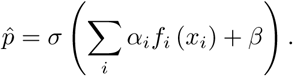

Here 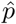 denotes the estimated probability that the input corresponds to a positive sample and *σ*(*x*) = (1 + *e*^*x*^)^*−*1^ denotes the sigmoid function. GAMs were traditionally fitted with splines or decision trees, but very recent work parameterised the functions *f*_*i*_ by simple multi-layer perceptrons giving rise to the so called Neural Additive Models (NAM) [16]. GAMs, and as a special case NAMs, are inherently interpretable because it is possible to visualise the individual functions *f*_*i*_ as a function of their corresponding feature. This allows users to globally interpret the model. Moreover, given a specific data sample *x*_*i*_, it is possible to analyse the individual importance of each feature by comparing the contribution, that is, the individual summands *α*_*i*_*f*_*i*_(*x*_*i*_) to the overall classification score.

Unlike classical GAMs, NAMs are not limited to tabular data. It is possible to choose *f*_*i*_ to be any type of neural network architecture and consequently input whole time series instead of tabular values. There exists little work that considers the GAM framework in this more general non-tabular setting. See for example [27] for an application in genomics. When using more general networks to input whole time series *x*_*i*_ instead of tabular features, the “global” interpretability of *f*_*i*_ is lost. Because *x*_*i*_ is now high dimensional, it is impossible to visualise *f*_*i*_ as a simple function of *x*_*i*_. We could, however, still read out the additive contribution of each network *α*_*i*_*f*_*i*_(*x*_*i*_) and therefore still quantify the importance of an individual time series modality to the overall classification. This analysis could either be done for a single specific input or over a whole set of inputs to gauge the overall signal modality importance.

### Activation Maps (Question 2)

To alleviate the loss of the global interpretability, we chose deep fully convolutional networks (FCNs) [28] as our functions *f*_*i*_. By parameterising the modality functions *f*_*i*_ as FCNs, we could achieve local interpretability by computing activation maps [29]. FCNs have been used extensively in time series classification [30]. They consist of several layers of intertwined operations of convolutional filters and non-linear activation functions, as well as a final output layer, which computes a global average over the convolutional filter activations across the temporal dimension.

The computed activation maps are inherently linked to the inductive bias of the FCN: The FCN detects temporally local patterns in the time series on the scale of the size of the used convolutional filters. As a result, activation maps are temporally local “explanations” that reflect the filter activations across the temporal dimension and thus allowed us to investigate which parts of the signal are discriminative for the classification (Question 2). Due to the linearity of the final global average operation, activation maps were easily integrated into the NAM framework: The additive contributions for each individual signal modality were just taken to be the average filter activation across time.

### Architecture and training details

We implemented the presented NAM, as well as all downstream analysis in Python using the Pytorch [31] and the Scikit-learn [32] library. The individual subnetworks consisted of three convolutional layers. Kernel sizes were selected with nested cross-validation and ranged between 5 and 17. We used zero padding to preserve the time dimension of each input signal. Like the kernel size, the number of convolutional hidden channels was also selected with nested cross validation, which resulted in 20 hidden channels per layer. After each convolutional layer, we applied batch normalisation [33] and the rectified linear units (ReLUs) [34]. Before the global average pooling layer, network activations were linearly combined per time point to produce one single time series activation map for each signal modality.

The Adam optimiser [35] was used to train the classifier with a standard binary cross entropy loss. We trained the networks for 10 epochs with a learning rate of 0.0001 and a weight decay of 0.001. These choices resulted in optimal performance in every validation fold. The previously mentioned network hyper-parameters were also stable across validation folds. The stability of hyperparameters across subjects was a consequence of our leave-one-out approach, where training sets differ only slightly between validation folds.

## Data Availability

All data produced are available online at

https://zenodo.org/record/7711137

## Data Availability

The datasets analysed during the current study are available on Zenodo: https://zenodo.org/record/7711137, (DOI: 10.5281/zenodo.7711137).

## Code Availability

The underlying code for this study is available in neonatal_apnea_prediction and can be accessed via this link https://github.com/mackelab/neonatal_apnea_prediction.

## Author contributions

J.V. developed the required model to process and analysis the data, wrote the first draft of the manuscript, and approved the final draft. K.L. included eligible infants, collected, and annotated data, and provided input to manuscript editing and approved the final draft. T.M.H.D., P.A.D. and O.K. provided intellectual input to the study, data interpretation, and manuscript editing, and approved the final draft. J.H.M. assisted in developing the required model to process the data, provided intellectual input to the study, data interpretation, and manuscript editing, and approved the final draft. C.F.P. conceived this project, obtained funding, co-ordinated study design and implementation of data collection and analysis tools, supervised the study, oversaw data analysis and interpretation, participated in manuscript editing, and approved the final draft.

## Acknowledgements

We would like to thank the staff, parents and infants involved in this study, Ms. Gabriele Hilber-Moessner who provided additional support in obtaining the data, and Dr. Mirja Quante who provided valuable advise with annotating the data. We also thank the International Max Planck Research School for Intelligent Systems (IMPRS-IS) and the AI4Med-BW graduate program for supporting J.V. This work was funded by the German Research Foundation (DFG) through Germany’s Excellence Strategy (EXC-Number 2064/1, Project number 390727645). The funder played no role in study design, data collection, analysis and interpretation of data, or the writing of this manuscript.

### Appendix

#### Significance analysis

To avoid confusion, we only reported the uncertainties over different patients in the main text. But for a more rigorous performance comparison between the Neural Additive Model (NAM) and single modality networks, we ran the complete training and testing pipeline ten times and compute the average AuROC and standard deviation over all patients (Table 1).

**Table 1:** NAM vs. single modality networks. Average AuROC and standard deviation of NAM and signal modality networks over 10 independent training and testing runs. The two-sample *t*-test is computed over *r* = 10 values per model after averaging over all patients. Conversely, the Wilcoxon tests are computed over *n* = 19 values per model with the performance for each patient averaged over all 10 runs. All tests are two-sided.

| Model | AuROC | SD | $t$-test | Wilcoxon |
| --- | --- | --- | --- | --- |
| NAM | 0.803 | 0.0077 | - | - |
| NP | 0.753 | 0.0067 | $< 0.001$ | $= 0.009$ |
| T+A | 0.735 | 0.0042 | $< 0.001$ | $< 0.001$ |
| SpO<sub>2</sub> | 0.733 | 0.0058 | $< 0.001$ | $= 0.006$ |
| PPG | 0.694 | 0.0074 | $< 0.001$ | $< 0.001$ |
| HR | 0.627 | 0.0073 | $< 0.001$ | $< 0.001$ |
| PCO<sub>2</sub> | 0.614 | 0.0077 | $< 0.001$ | $< 0.001$ |

We then performed a significance analysis over the differences in average performance between the NAM and the single modality networks. Two statistical tests are computed: Wilcoxon signed-rank tests over the *n* = 19 patients and two-sample *t*-tests over the *r* = 10 runs (Table 1).

We complete the significance analysis by comparing all pairs of performances and additive contributions (Table 2 and Table 3) again using Wilcoxon signed-rank tests.

**Table 2:** Statistical test results for single modality network performances. The Wilcoxon tests are computed over *n* = 19 individual AuROC performances where the performance value for each patient is the average over all 10 runs. All tests are two-sided.

| Model | NP | T+A | SpO<sub>2</sub>, | PPG | HR |
| --- | --- | --- | --- | --- | --- |
| T+A | $= 0.032$ | | | | |
| SpO<sub>2</sub> | $= 0.241$ | $= 0.829$ | | | |
| PPG | $= 0.011$ | $= 0.087$ | $= 0.568$ | | |
| HR | $= 0.007$ | $= 0.016$ | $= 0.005$ | $= 0.169$ | |
| PCO<sub>2</sub> | $= 0.012$ | $= 0.018$ | $= 0.003$ | $= 0.123$ | $= 0.49$ |

**Table 3:** Statistical test results for the additive contributions. The Wilcoxon tests are computed over *n* = 19 standard deviations of the patient-based additive contributions. The values for each patient are obtained by averaging over all 10 runs. All tests are two-sided.

| Model | NP | T+A | SpO<sub>2</sub>, | PPG | HR |
| --- | --- | --- | --- | --- | --- |
| T+A | $< 0.001$ | | | | |
| SpO<sub>2</sub> | $< 0.001$ | $= 0.679$ | | | |
| PPG | $< 0.001$ | $< 0.001$ | $= 0.005$ | | |
| HR | $< 0.001$ | $< 0.001$ | $< 0.001$ | $< 0.001$ | |
| PCO<sub>2</sub> | $< 0.001$ | $< 0.001$ | $< 0.001$ | $< 0.001$ | $= 0.275$ |

Table 4 shows the results of the permutation tests [18] for all patients for both the NAM and all single modality networks.

**Table 4:** Permutation tests for *m* = 1024 permutations for both the NAM and all single modality networks. Exact *p*-values are given for values larger then 0.001. The patient ordering does not correspond to the one in the main text.

| ID | NAM | Np | Th | SpO<sub>2</sub> | PPG | HR | PCO<sub>2</sub> |
| --- | --- | --- | --- | --- | --- | --- | --- |
| 01 | 0.001 | 0.001 | 0.001 | 0.001 | 0.001 | 0.001 | 0.017 |
| 02 | 0.001 | 0.001 | 0.001 | 0.001 | 0.001 | 0.001 | 0.001 |
| 03 | 0.001 | 0.001 | 0.002 | 0.001 | 0.001 | 0.001 | 0.001 |
| 04 | 0.001 | 0.001 | 0.005 | 0.062 | 0.008 | 0.002 | 0.001 |
| 05 | 0.001 | 0.001 | 0.001 | 0.001 | 0.008 | 0.374 | 0.983 |
| 06 | 0.001 | 0.001 | 0.001 | 0.001 | 0.001 | 0.607 | 0.001 |
| 07 | 0.001 | 0.001 | 0.001 | 0.001 | 0.001 | 0.044 | 0.921 |
| 08 | 0.001 | 0.001 | 0.001 | 0.001 | 0.001 | 0.037 | 0.998 |
| 09 | 0.001 | 0.03 | 0.02 | 0.001 | 0.2 | 0.027 | 0.005 |
| 10 | 0.001 | 0.001 | 0.001 | 0.001 | 0.001 | 0.997 | 0.7 |
| 11 | 0.004 | 0.845 | 0.001 | 0.001 | 0.727 | 0.001 | 0.003 |
| 12 | 0.001 | 0.001 | 0.001 | 0.001 | 0.008 | 0.001 | 0.001 |
| 13 | 0.001 | 0.001 | 0.001 | 0.001 | 0.001 | 0.001 | 0.001 |
| 14 | 0.001 | 0.001 | 0.001 | 0.001 | 0.001 | 0.001 | 0.07 |
| 15 | 0.001 | 0.001 | 0.001 | 0.006 | 0.008 | 0.016 | 0.006 |
| 16 | 0.001 | 0.001 | 0.001 | 0.001 | 0.001 | 0.366 | 0.001 |
| 17 | 0.121 | 0.157 | 0.558 | 0.713 | 0.103 | 0.031 | 0.004 |
| 18 | 0.001 | 0.001 | 0.001 | 0.001 | 0.001 | 0.853 | 0.778 |
| 19 | 0.001 | 0.001 | 0.014 | 0.001 | 0.661 | 0.001 | 0.001 |

#### Description and definition of adverse events

The criteria used to annotate the recorded polygraphy signals are given in the following. More details can be found in [17].

1. Obstructive apnea
  - Lasts for ≥ 2 breaths with respiratory effort present on inductance belt
2. Central apnea
  - Lasts for ≥ 2 breaths with associated SpO2 decrease by ≥ 3%
  - Lasts for ≥ 20 seconds
  - Arousal or HR *<* 50bpm for ≥ 5 seconds or *<* 60bpm for *>* 15 seconds
3. Mixed apnea
  - Lasts for ≥ 2 breaths with component of both obstructive and central apnea
4. Obstructive hypopnea
  - Nasal flow ≤ 70% of baseline for ≥ 2 breaths with associated SpO2 decrease by ≥ 3%
  - Arousal
  - Hypopnea with increased inspiratory flattening
  - Thoracoabdominal paradox
5. Central hypopnea
  - Without above-mentioned features of obstruction
6. Hypoxia
  - SpO2 decrease by ≥ 3% within a 5 seconds duration
7. Movement
  - Gross movement observed for at least 15 seconds
  - Eye opening
  - Two episodes of movement need to be separated by at least 15 seconds

## Additional activation maps

Here, we provide the activation maps of all six signal modalities of the examples shown in the main text as well as additional examples.

**Figure 4:**
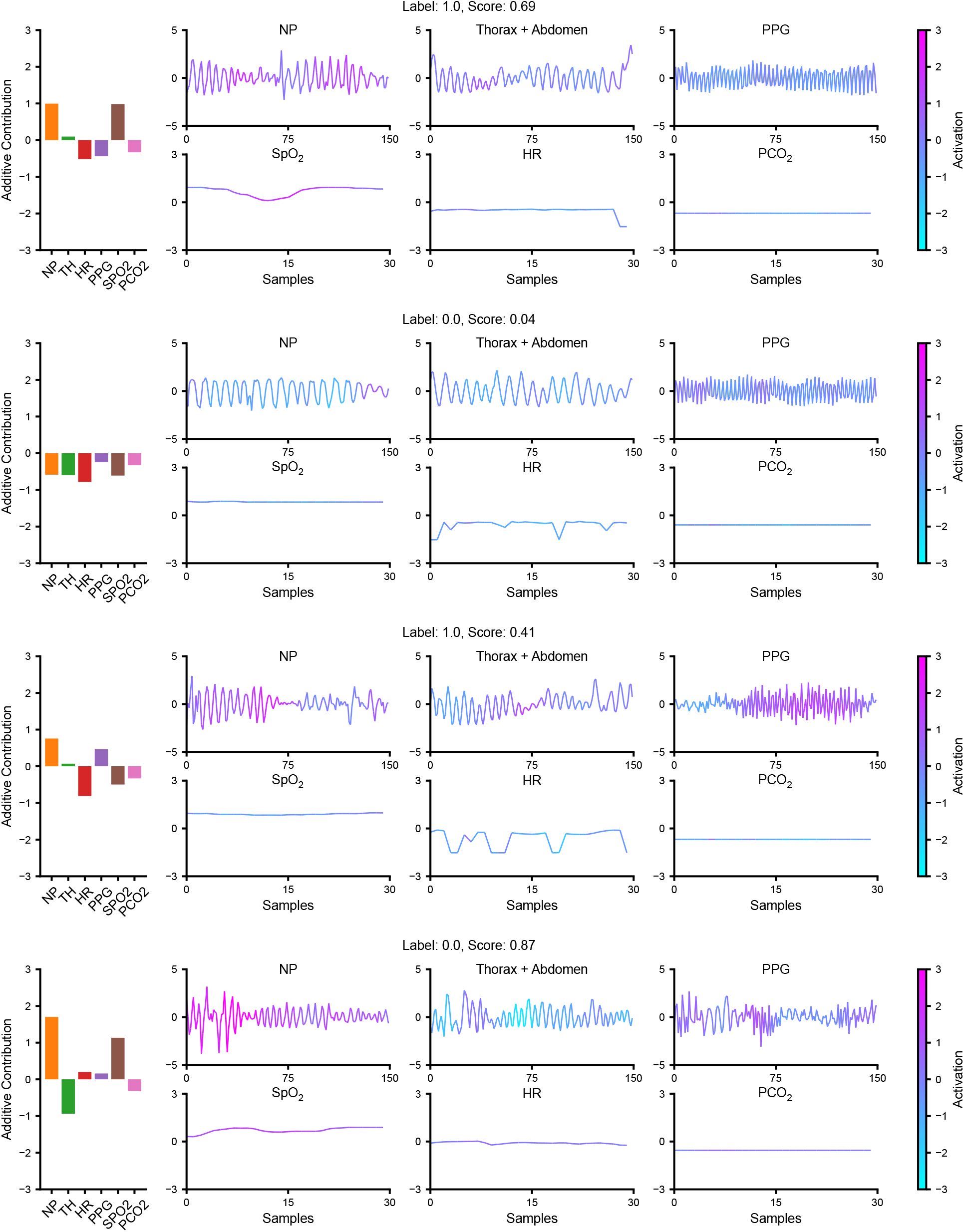
Full activations maps for the examples shown in the main text.

**Figure 5:**
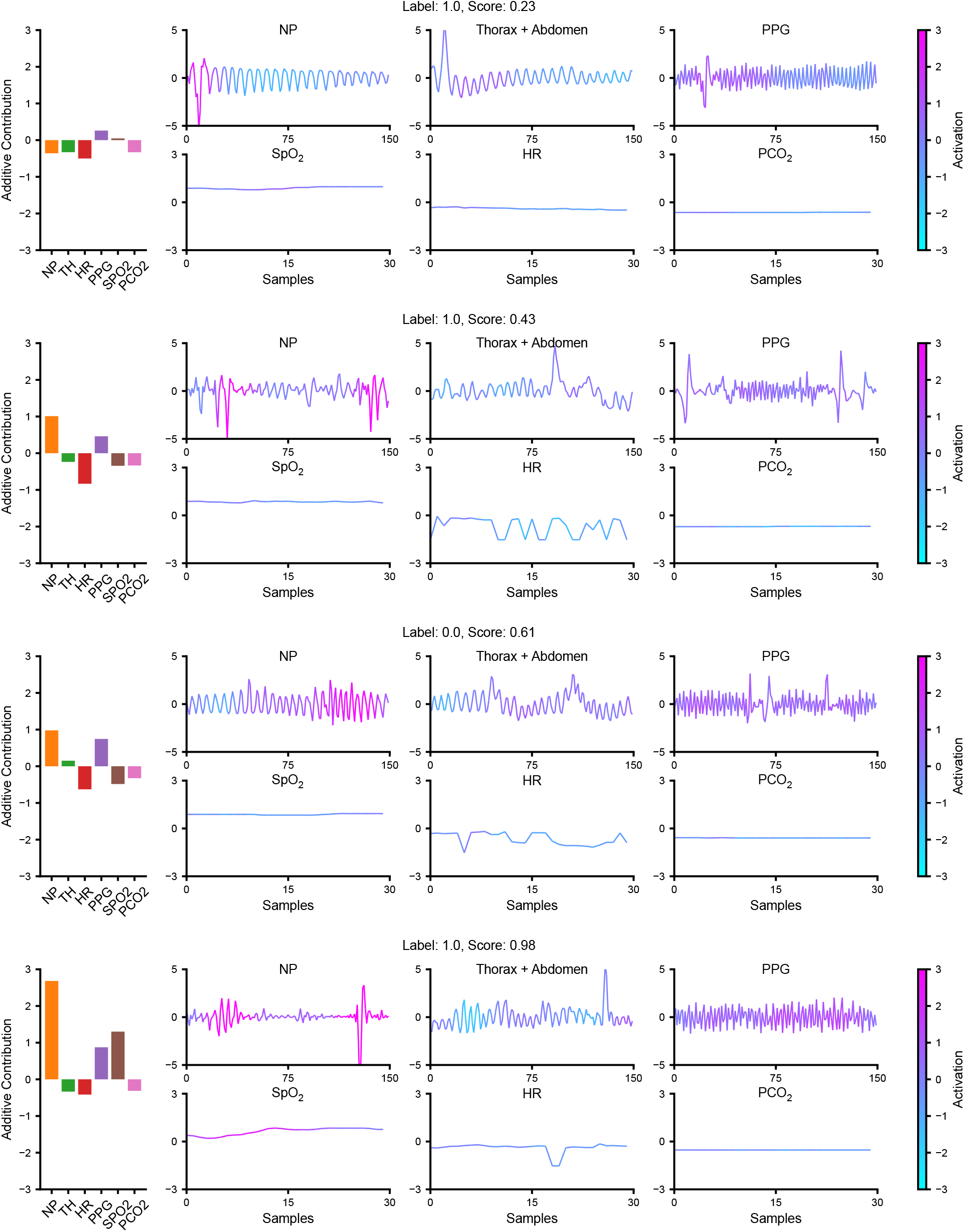
Additional activation maps.

**Figure 6:**
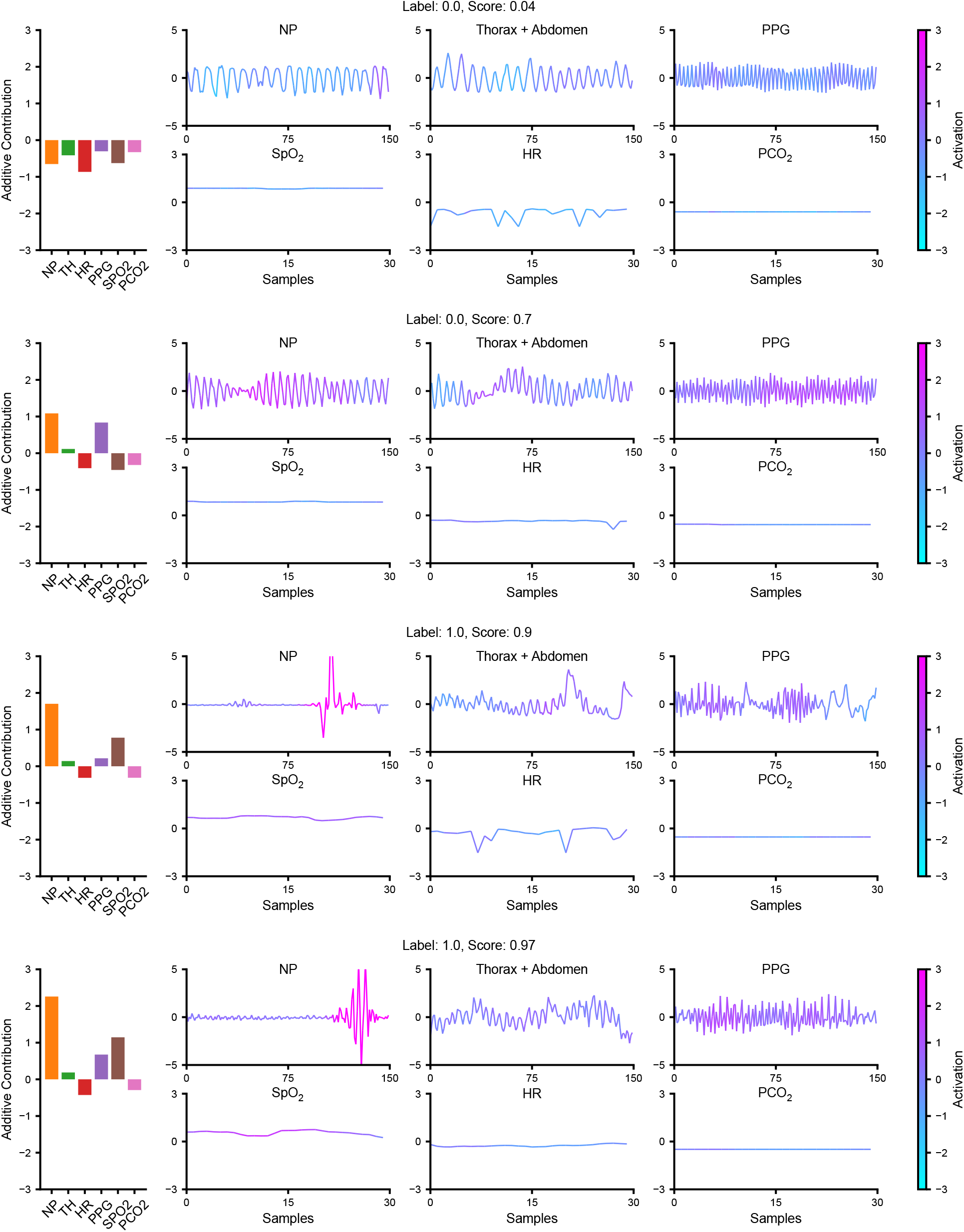
Additional activation maps.

## References

[1] Bloch-Salisbury, E., Indic, P., Bednarek, F. & Paydarfar, D. Stabilizing immature breathing patterns of preterm infants using stochastic mechanosensory stimulation. Journal of Applied Physiology 107, 1017–1027 (2009).

[2] Cattani, L. et al. Monitoring infants by automatic video processing: A unified approach to motion analysis. Computers in Biology and Medicine 80, 158–165 (2017).

[3] Vergales, B. D. et al. Accurate automated apnea analysis in preterm infants. American journal of Perinatology 31, 157–162 (2014).

[4] Clark, M. T. et al. Breath-by-breath analysis of cardiorespiratory interaction for quantifying developmental maturity in premature infants. Journal of Applied Physiology 112, 859–867 (2012).

[5] Altuve, M., Carrault, G., Beuchee, A., Pladys, P. & Hernández, A. I. Online apnea-bradycardia detection using hidden semi-Markov models. In Annual International Conference of the IEEE Engineering in Medicine and Biology Society, 4374–4377 (IEEE, 2011).

[6] Zuzarte, I., Sternad, D. & Paydarfar, D. Predicting apneic events in preterm infants using cardio-respiratory and movement features. Computer Methods and Programs in Biomedicine 209, 106321 (2021).

[7] Pravisani, G., Beuchee, A., Mainardi, L. & Carrault, G. Short term prediction of severe bradycardia in premature newborns. In Computers in Cardiology, 725–728 (IEEE, 2003).

[8] Mahmud, M. S., Wang, H. & Kim, Y. Accelerated prediction of bradycardia in preterm infants using time-frequency analysis. In International Conference on Computing, Networking and Communications (ICNC), 468–472 (IEEE, 2019).

[9] Shirwaikar, R. D., Acharya, D., Makkithaya, K., Surulivelrajan, M. & Srivastava, S. Optimizing neural networks for medical data sets: A case study on neonatal apnea prediction. Artificial intelligence in medicine 98, 59–76 (2019).

[10] Lim, K. et al. Predicting apnoeic events in preterm infants. In Frontiers in Pediatrics, 570 (2020).

[11] Ismail Fawaz, H., Forestier, G., Weber, J., Idoumghar, L. & Muller, P.-A. Deep learning for time series classification: A review. Data mining and knowledge discovery 33, 917–963 (2019).

[12] Futoma, J. et al. An improved multi-output gaussian process RNN with real-time validation for early sepsis detection. In Machine Learning for Healthcare Conference, 243–254 (PMLR, 2017).

[13] Lea, C., Flynn, M. D., Vidal, R., Reiter, A. & Hager, G. D. Temporal convolutional networks for action segmentation and detection. In Proceedings of the IEEE Conference on Computer Vision and Pattern Recognition, 156–165 (2017).

[14] Rudin, C. Stop explaining black box machine learning models for high stakes decisions and use interpretable models instead. Nature Machine Intelligence 1, 206–215 (2019).

[15] Semenova, L., Rudin, C. & Parr, R. On the existence of simpler machine learning models. In 2022 ACM Conference on Fairness, Accountability, and Transparency, 1827–1858 (2022).

[16] Agarwal, R. et al. Neural additive models: Interpretable machine learning with neural nets. In Advances in Neural Information Processing Systems, vol. 34, 4699–4711 (2021).

[17] Lim, K. et al. Should obstructive hypopneas be included when analyzing sleep studies in infants with Robin sequence? Sleep Medicine 98, 9–12 (2022).

[18] Ojala, M. & Garriga, G. C. Permutation tests for studying classifier performance. Journal of Machine Learning Research 11, 1833–1863 (2010).

[19] Lou, Y., Caruana, R., Gehrke, J. & Hooker, G. Accurate intelligible models with pairwise interactions. In Proceedings of the 19th ACM SIGKDD international conference on Knowledge discovery and data mining, 623–631 (2013).

[20] Cawley, G. C. & Talbot, N. L. C. On over-fitting in model selection and subsequent selection bias in performance evaluation. Journal of Machine Learning Research 11, 2079–2107 (2010).

[21] Esquer, C., Claure, N., D’Ugard, C., Wada, Y. & Bancalari, E. Mechanisms of hypoxemia episodes in spontaneously breathing preterm infants after mechanical ventilation. Neonatology 94, 100–104 (2008).

[22] Di Fiore, J. M. Neonatal cardiorespiratory monitoring techniques. Seminars in Neonatology 9, 195–203 (2004).

[23] Richards, J. M. et al. Sequential 22-hour profiles of breathing patterns and heart rate in 110 full-term infants during their first 6 months of life. Pediatrics 74, 763–777 (1984).

[24] Kelly, D. H. & Shannon, D. C. Treatment of apnea and excessive periodic breathing in the full-term infant. Pediatrics 68, 183–186 (1981).

[25] Hastie, T. J. Generalized Additive Models, 249–307 (Routledge, 2017).

[26] Lou, Y., Caruana, R. & Gehrke, J. Intelligible models for classification and regression. In Proceedings of the 18th ACM SIGKDD international conference on Knowledge discovery and data mining, 150–158 (2012).

[27] Srivastava, D., Aydin, B., Mazzoni, E. O. & Mahony, S. An interpretable bimodal neural network characterizes the sequence and preexisting chromatin predictors of induced transcription factor binding. Genome biology 22, 1–25 (2021).

[28] Gu, J. et al. Recent advances in convolutional neural networks. Pattern recognition 77, 354–377 (2018).

[29] Zhou, B., Khosla, A., Lapedriza, A., Oliva, A. & Torralba, A. Learning deep features for discriminative localization. In Proceedings of the IEEE conference on computer vision and pattern recognition, 2921–2929 (2016).

[30] Wang, Z., Yan, W. & Oates, T. Time series classification from scratch with deep neural networks: A strong baseline. In 2017 International joint conference on neural networks (IJCNN), 1578–1585 (IEEE, 2017).

[31] Paszke, A. et al. Pytorch: An imperative style, high-performance deep learning library. In Advances in neural information processing systems, vol. 32 (2019).

[32] Pedregosa, F. et al. Scikit-learn: Machine learning in Python. Journal of Machine Learning Research 12, 2825–2830 (2011).

[33] Ioffe, S. & Szegedy, C. Batch normalization: Accelerating deep network training by reducing internal covariate shift. In International conference on machine learning, 448–456 (PMLR, 2015).

[34] Arora, R., Basu, A., Mianjy, P. & Mukherjee, A. Understanding deep neural networks with rectified linear units. In arXiv preprint arXiv:1611.01491 (2016).

[35] Kingma, D. P. & Ba, J. Adam: A method for stochastic optimization. In arXiv preprint arXiv:1412.6980 (2014).

